# PUBLIC ATTITUDES TOWARDS FOOD TAXES FOR HEALTHY EATING IN THE UNITED KINGDOM

**DOI:** 10.1101/2024.10.15.24315520

**Authors:** Mario Martínez-Jiménez, Hannah Brinsden, Franco Sassi

**Affiliations:** Centre for Health Economics & Policy Innovation, Department of Economics & Public Policy, Imperial College Business School, London, UK; The Food Foundation, London, UK

**Keywords:** Fiscal policies, Public attitudes, Food policy, Taxes, Obesity

## Abstract

**Introduction:** Appropriately designed food taxes can improve diet quality and health. Fiscal levers are used in several countries to combat the rise in obesity and diet-related diseases. This study aims to investigate public attitudes, knowledge, and policy preferences regarding food taxes for promoting healthy eating in the UK.

**Methods:** A survey was administered through YouGov Plc to a nationally representative sample of 2,125 adults, gathering information on: acceptability and support for different types of food taxes, awareness and knowledge of existing taxes, and preferences for the characteristics of possible new taxes.

**Results:** Overall, 48% of respondents support higher taxes on unhealthy foods, rising to 72% if taxes made healthy foods more affordable. Respondents with high socio-economic status and those living in London showed the highest support. Respondents had limited awareness of existing food and beverage taxes, and prioritised discretionary items such as cakes and crisps for possible increased taxation.

**Conclusions:** The survey shows a high level of support for taxing unhealthy foods, as well as concern for the affordability of healthy foods. A carefully designed holistic approach to food taxation can be part of a wider public health strategy and can be favourably met by the general population in the UK.

## 1. INTRODUCTION

The use of fiscal incentives to promote healthy eating has increased in the past decade, amid a continuing growth in obesity and diet-related diseases. While many countries currently apply taxes on sugar-sweetened beverages (Hattersley et al., 2023), a smaller number of countries have utilised taxes to promote healthy eating, and most of the latter are limited in scope (WHO, 2022). This paper aims to provide insights into public attitudes for policymakers seeking to use fiscal levers to address the challenge of unhealthy dietary choices.

Food taxation has the potential to improve health by reducing the consumption of unhealthy foods and improving diet quality (Gorski et al., 2015). The ultimate goal of health taxes applied on food is to curb diet-related conditions such as obesity, diabetes and tooth decay (Griffith et al., 2021; Colombo et al., 2023; Hashem et al., 2024).

Public support for taxes is an important factor for successfully getting a tax implemented (Thow et al., 2022), even more so in the context of the rising cost of living where many people are concerned about food prices (Broadbent et al., 2023). Understanding public attitudes towards food taxes is essential for the successful implementation and sustainability of such health policies. This study aims to provide valuable information for policymakers, enabling them to design and advocate for food taxation measures that are not only effective in promoting healthier eating habits but also supported and acknowledged by the public. This, in turn, can lead to improved public health outcomes and a reduction in diet-related diseases in the UK.

Our study explores public attitudes towards the implementation of food taxes aimed at promoting healthy eating by conducting a nationally representative survey of 2,125 adults in the UK between April 12th and 14th, 2024. Our survey covers three main areas: (i) support, in principle, for a new tax on unhealthy food; (ii) awareness of the current taxes on foods and non-alcoholic beverages in the UK, the Value-Added Tax (VAT) and the Soft Drink Industry Levy (SDIL), and (iii) public preferences regarding specific characteristics of a possible new tax on unhealthy foods. Our headline results show that there is support for food taxes on unhealthy food, particularly when coupled with measures to make healthier food more affordable. Findings also show poor awareness of existing taxes, which is poorer for the SDIL than for VAT. Support is higher among females, people of high social class, and people living in London. Priority targets for possible taxes on unhealthy foods included food groups such as cakes, potato crisps, hot takeaways and ready meals.

### Literature review

Food taxation has been extensively studied as a potential public health measure aimed at incentivising healthier dietary choices. Research indicates that prices and promotions within the food environment can significantly influence consumer behaviour, often encouraging the selection of foods contributing to less healthy diets. Taxation of unhealthy products, such as tobacco, alcohol or sugar, is not new, but traditionally these taxes have been used for purely fiscal reasons, i.e., to generate tax revenues in order to finance public spending.

Empirical evidence from various countries demonstrates that taxation can be an effective tool for promoting healthier dietary choices (Andreyeva et al., 2022; Pineda et al., 2024). Scarborough et al. (2020) show that in 2016, as a result of the SDIL — a tax on soft drinks that contain more than 5 g of sugar per 100 mL – the percentage of drinks with sugar fell from 49% to 15% between September 2015 and February 2019 in the United Kingdom. Taillie et al. (2017) show a 12% reduction in sales in the first year following the implementation of Mexico’s 8% nonessential energy-dense food tax, driven by lower-income households. Similarly, Berkeley’s sugar-sweetened beverages (SSB) tax resulted in a 21% drop in the consumption of sugary drinks, demonstrating the potential of such taxes to alter consumer behaviour. In 2011, Denmark implemented a €2.14 per kg tax on saturated fat for products with more than 2.3 g per 100 g of saturated fat. Although this tax was subsequently repealed in 2013, for the duration of implementation, saturated fat purchases were reduced by 4%, and deaths attributable to non-communicable diseases were estimated to have been reduced by 0.4% (Smed et al., 2016). Hungary has introduced a tax targeting prepacked foods that are high in salt, sugar, or caffeine (at varying tax rates), which has been associated with a 3.4% reduction in the consumption of processed food (and a compensatory 1.1% increase in unprocessed food) as shown by Bíró et al. (2015).

Public attitudes towards taxes, particularly those aimed at improving health, vary widely depending on the context. In the case of health taxes, such as those on tobacco (Filippidis et al., 2014; Farley et al., 2015) and sugary drinks (Jou et al., 2014; Miller et al., 2019; Pell et al., 2019; Chriqui et al., 2020), there tends to be a higher level of support when the public perceives a clear link between the tax and health benefits. In contrast, general taxes are often viewed through a lens of economic impact and fairness (Sumino, 2016; Lachapelle, 2021). Public trust in government plays a significant role in shaping these attitudes; higher trust correlates with greater acceptance of both specific and general taxes. Eykelenboom et al. (2019) conducted a systematic review synthesising the existing literature on the political and public acceptability of an SSB tax covering countries around the world (e.g., the US, Australia, the UK, Mexico, China, France, New Zealand, among others). The authors found that of the public, 42% supported a tax on SSB, 39% supported an SSB tax as a means of tackling obesity, and 66% supported such a tax where the revenue was used “appropriately,” e.g. for health initiatives.

Regarding the acceptability of food taxes, Mazzocchi et al. (2015) found that the main drivers of policy support in five European countries are attitudinal factors, particularly the belief that obesity is largely due to the excessive availability of unhealthy foods. Socio-demographic characteristics and political preferences, however, are not strongly correlated with support for these policies. In Germany, Jurkenbeck et al. (2020) found that a large majority of citizens accept nutrition policy interventions, mostly among those individuals who generally maintain a healthy diet and those who struggle with dietary habits. In Norway, Grimsrud et al. (2019) demonstrated that there is significant public acceptance and willingness to pay for cost-effective taxes, particularly on red meat, despite the public’s scepticism towards the introduction or increase of taxes aimed at environmental improvement. This suggests that while environmental taxes may face resistance, targeted nutritional interventions addressing public health concerns can receive substantial support across diverse demographic groups.

## 2. MATERIALS AND METHODS

### 2.1. Survey data collection and sample

We collected our survey data between April 12th and 14th, 2024 through YouGov Plc (https://yougov.co.uk/). The survey was conducted using online interviews with members of the YouGov panel, which consists of 185,000+ individuals who have agreed to participate in the Company’s surveys. To ensure the sample is representative of the UK population or specific sub-groups, stratified sampling techniques are used to select respondents based on key demographics such as age, gender, region, and socio-economic status. Surveys are distributed to the selected sample via email invitations or through the YouGov platform, allowing respondents to respond at their convenience. YouGov Plc typically achieves a response rate of between 35% and 50% for surveys, although this can vary depending on the subject matter, complexity, and length of the questionnaire.

The responding sample is weighted to match the profile of the original target sample. During the survey period, response rates and data quality are monitored, and if certain demographics are underrepresented, additional invitations may be sent to those groups. The survey was completed by 2,125 adults between

April 12th and 14th, 2024, using an online questionnaire. Table 1 shows that the resulting (weighted) sample is representative of the UK population. One dimension in which our sample may differ is regarding political preferences (captured by the 2019 General elections and EU referendum in 2016), in which missing observations account for around 26 and 23 per cent of our sample (see Appendix A). For that reason, political party preference is not used in our main analysis. All analyses reported in this paper are using sample weights.

**Table 1.** Sample representativeness and summary statistics of survey participants.

|  |  | Weighted Sample |  | Unweighted Sample |  | Population % |
| --- | --- | --- | --- | --- | --- | --- |
|  |  | N | % | N | % |  |
| **Gender** | *Male* | 1031 | 48.5 | 966 | 45.5 | 49.45 |
|  | *Female* | 1094 | 51.5 | 1159 | 54.5 | 50.55 |
| **Age** | *18-24* | 223 | 10.5 | 192 | 9 | 8.3 |
|  | *25-49* | 878 | 41.3 | 894 | 42.1 | 32.9 |
|  | *50-64* | 527 | 24.8 | 534 | 25.1 | 19.5 |
|  | *65+* | 497 | 23.4 | 505 | 23.8 | 18.4 |
| **Social Grade** | *ABC1* | 1211 | 57 | 1258 | 59.2 | 57 |
|  | *C2DE* | 914 | 43 | 867 | 40.8 | 43 |
| **Country** | *England* | 1785 | 84 | 1790 | 84.2 | 84 |
|  | *Wales* | 104 | 4.9 | 113 | 5.3 | 4.7 |
|  | *Scotland* | 181 | 8.5 | 172 | 8.1 | 8.2 |
|  | *Northern Ireland* | 55 | 2.6 | 50 | 2.4 | 2.9 |
| **Region in England** | *North* | 499 | 27.9 | 507 | 28.3 | 27 |
|  | *Midlands* | 342 | 19.2 | 354 | 19.8 | 20 |
|  | *London* | 251 | 14.1 | 217 | 12.1 | 16 |
|  | *South* | 693 | 38.9 | 712 | 39.8 | 32 |
| **Total** |  | **2,125** | **100** | **2,125** | **100** | **100** |

*Notes:* The total sample size is 2,125 adults. Fieldwork was undertaken between 12th and 14th April 2024. The survey was carried out online. The figures have been weighted and are representative of all UK adults (aged 18+). *Source:* YouGov and Office National Statistics (ONS).

### 2.2. The questionnaire and demographic characteristics

The questionnaire is structured in three parts, described as *(i)* acceptability/support, *(ii)* awareness/knowledge and *(iii)* preferences on characteristics of a possible food tax. Our questionnaire includes eight closed-ended questions. Three questions were asked about awareness/knowledge of the UK tax system for food and drinks (VAT and SDIL), and two questions addressed the acceptability of taxes on unhealthy food. Finally, a set of questions captures individuals’ preference for a possible new tax on unhealthy foods. This refers to one question focused on which food groups should or should not have a higher tax, while two more questions asked about specific goals, such as discouraging people from purchasing certain products and encouraging them to buy others instead.

Based on previous public opinion research on perception and attitudes towards taxes and policies, we asked respondents about their basic socioeconomic and demographic characteristics, including their age (<30; 30-44; 45-64; >64), social class (“Grade A: Professionals; very senior managers business; top-level civil servants”, “Grade B: Middle-management executives/Principal officers/Top management or owners of small business” or “Grade C1: Junior management/varied responsibilities and educational requirement”, and zero if “Grade C2: Skilled manual workers/Manual workers with responsibility for other people”, “Grade D: Semi-skilled and unskilled manual workers, apprentices and trainees of skilled workers” or “Grade E: Long-term recipients of state benefits/Unemployed/Off sick/casual workers”. We also controlled for region and country (London, Scotland, Wales, North Ireland and England). Appendix C provides the full questionnaire as well as links to each country’s questionnaire in the original language.

Our study measures respondents’ support for food taxes that promote healthy eating through two main questions: “Would you support or oppose a higher tax on unhealthy foods?” and “Would you support a higher tax on unhealthy foods if money makes healthy food cheaper?”

### 2.3. Statistical approach

To investigate the relationship between respondent characteristics and support for food taxes towards healthy eating habits, we estimate a linear probability model (LPM). A set of outcome variables captures to what extent a tax on unhealthy foods is supported by respondents. These variables are binary regarding the extent of support for taxes on unhealthy food, taking values one if the respondent “strongly/somewhat supports” and zero if the respondent “strongly/somewhat opposes’. It is important to note that “don’t know” observations are not accounted for in the main statistical analysis. This means that the statistical analysis focuses only on the responses where a clear opinion was provided, providing a clearer picture of the attitudes and preferences of the respondents who expressed a definite opinion. Sociodemographic characteristics included in the analysis are age groups, a binary variable capturing manual/non-manual social class (which takes one if the respondent’s social class grade is A/B/C1 and zero if C2/D/E. We are also controlling for regions/countries. We provide estimates with robust standard errors, and survey weights are used in all the regressions. Robustness checks are performed, including using an alternative statistical model (logistic regression), controlling for political preferences, and testing for multicollinearity bias when selecting our control variables. Results for all the robustness checks are available in Appendix B.

## 3. RESULTS

### 3.1. Knowledge of existing taxes applied to food and non-alcoholic drink products

We first analyse the level of awareness of existing taxes applied to foods and non-alcoholic beverages, VAT and SDIL. We found greater awareness of VAT compared to SDIL, with just 4% considered to have ‘little knowledge’ of VAT, compared to 24.3% for SDIL. There was, however, a similar number of people who we classify as ‘very knowledgeable’ across both taxes (27.4% for VAT and 25.6% for SDIL) – see Figure A1 in Appendix A.

We then asked whether a selection of foods have or do not have SDIL and VAT applied to them. While around 68% correctly identified that soft drinks have VAT, only 28.9% identified that bottled water had VAT applied. There is also high awareness amongst respondents that foods from both takeaways (69%) and restaurants (78.8%) have VAT applied. More than half of respondents (57.5%) incorrectly thought that cakes had VAT applied, with only 18.9% correctly identifying that they did not. The responses on the two meat options (fresh and processed) were mixed, with more people thinking VAT is applied to processed meat compared to fresh meat, around 40% correctly identifying that fresh meat does not have VAT, and only 24.4% correctly identifying that processed meat does not. Between 14% and 30% of respondents reported they do not know whether taxes apply for each of the mentioned food groups, with the highest rate for bottled water and the lowest rate for restaurant meals – see Figure A2 in Appendix A.

### 3.2. Support and opposition for higher food taxes for healthier eating habits

Overall, 48% of respondents supported higher food taxes (compared to 44% who opposed them), and this number would increase to 72% if the money raised were used to make healthier food more affordable. Only 23% opposed higher taxes on unhealthy foods if the revenue generated was used to make healthier food cheaper.

Regarding respondents’ level of support of food taxes on unhealthy food by sociodemographic characteristics, we find no substantial variation in support by age group, 18-24 (49 vs 42%). 25-49 (46 vs 45), 50-64 (50 vs 45). 65+ (50 vs 44), as we can see in Figure A3 in Appendix A. However, some differences are found by socioeconomic status. In Figure 1, we can see that people in non-manual professions are more likely to support higher taxes on unhealthy food, with percentage ranges of support between 60% and 48%. In contrast, those individuals who are long-term recipients of state benefits, unemployed or casual workers show the lowest support for these taxes, with around 34% of respondents supporting a higher tax on unhealthy foods.

**Figure 1.**
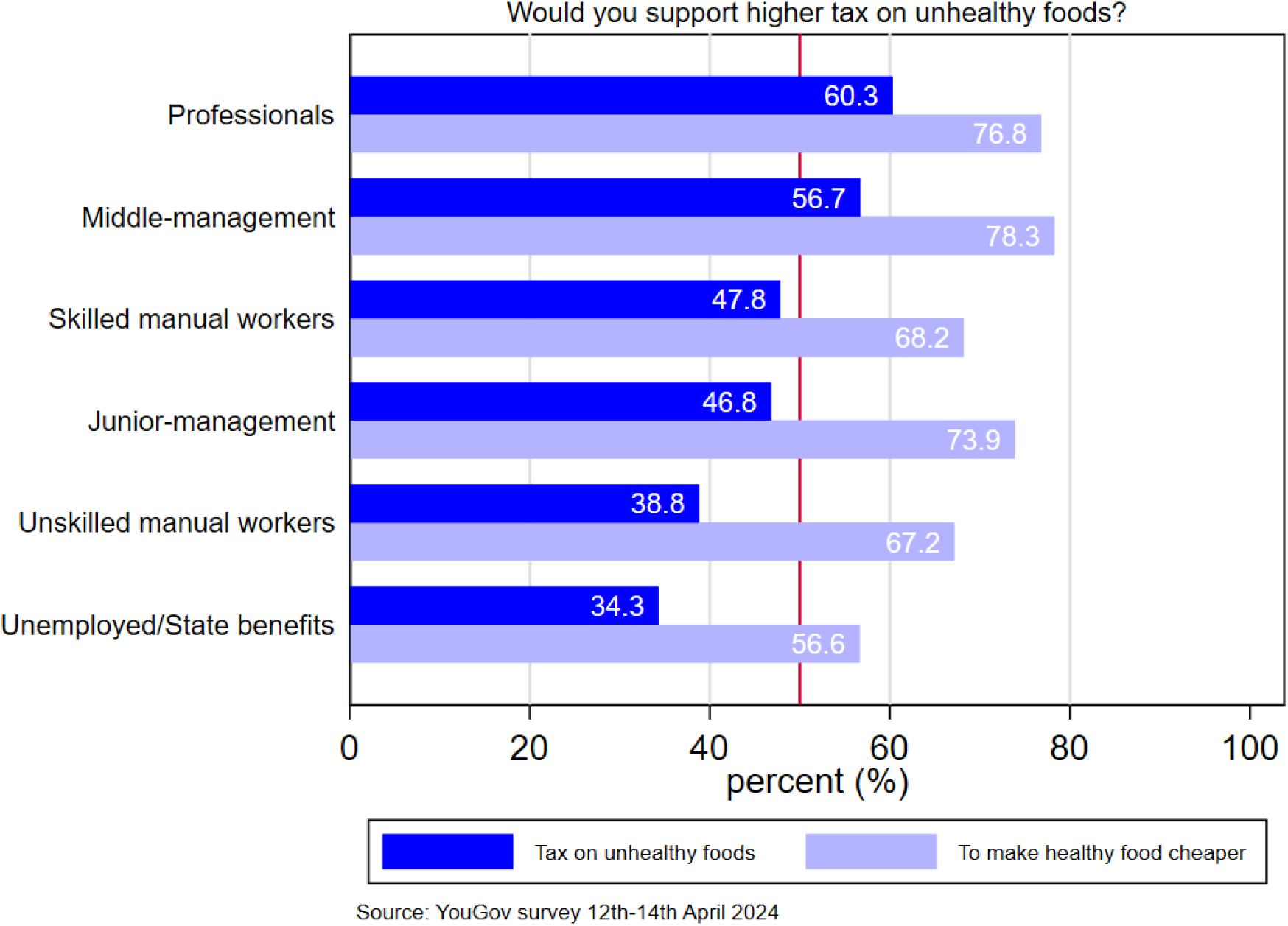
Share of respondents who agree (somewhat too strongly) that “Would you support a higher tax on unhealthy foods?” or “Would you support a higher tax on unhealthy foods to make healthy food cheaper” by socioeconomic status. *Notes*: The number of observations is 2,125; the base category includes the following responses: Strongly/somewhat opposed/Don’t know. The support category includes responses that Strongly/somewhat support. Socioeconomic status groups include the following professions: Professionals (grade A: Professionals; very senior managers in business; top-level civil servants); Middle-management (Grade B: Middle-management executives/Principal officers/Top management or owners of small businesses); Junior-management (Grade C1:Junior management/varied responsibilities and educational requirements); Skilled manual workers (Grade C2:Skilled manual workers/Manual workers with responsibility for other people); Unskilled manual workers (Grade D: Semi-skilled and unskilled manual workers, apprentices and trainees of skilled workers); and Unemployed/State benefits (Grade E: Long-term recipients of state benefits/Unemployed/Off sick/casual workers). Sample weights are used.

By region and country, we find the highest support in London, with around 54% of respondents supporting a tax on unhealthy food, increasing to 76.7% if the tax was used to make healthier food more affordable. Respondents living in Wales had the lowest support for the tax at 37.8%. Northern Ireland had the biggest increase in support when asked about making healthier food more affordable, with an increase from around 44% to 74%. The lowest support in England was among those living in the North East and West Midlands, with 44.6% and 44.5% reporting to support a tax on unhealthy food, respectively (see Figure A4 in Appendix A).

Figure 2 shows the results of the linear probability model of supporting taxes on unhealthy foods conditional to certain socioeconomic characteristics. In particular, we control the support on food taxes on age groups (reference group is those between 18-30 years old), gender, non-manual social class and living in London. Here, we can see that those in non-manual professions and living in London are statistically significantly more likely to support taxes on unhealthy foods compared to their counterparts. Similarly, women and those in non-manual professions are more likely to support a tax on unhealthy food that makes healthy food cheaper.

**Figure 2.**
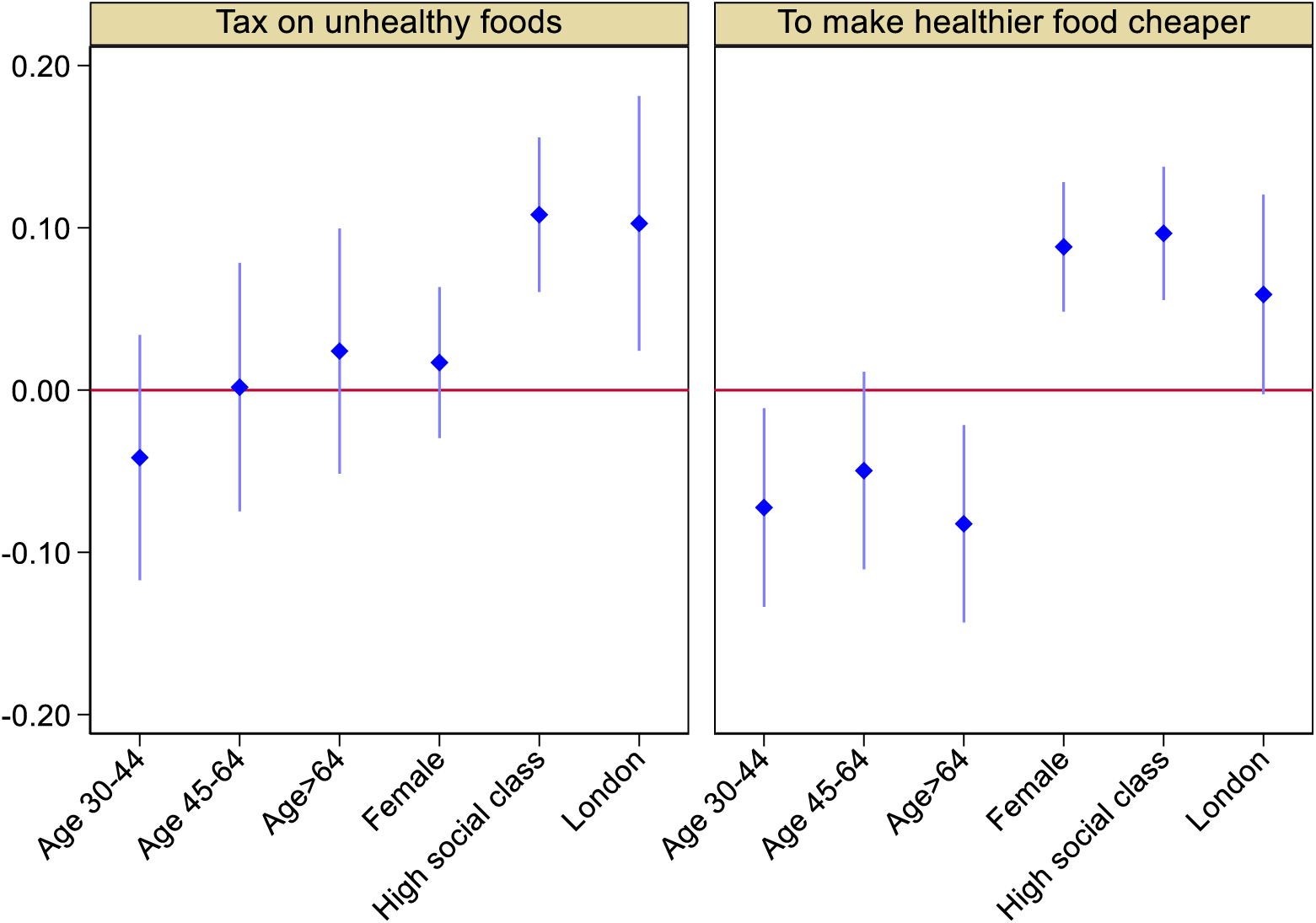
Correlation between support for a tax on unhealthy foods and socioeconomic characteristics. Notes: Base categories: age between 18-30; male; manual professions (social class grade: C2/D/E); other countries and English regions of the UK. The number of observations is 1,971, as those answering ‘do not know’ are not included.

**Figure 3.**
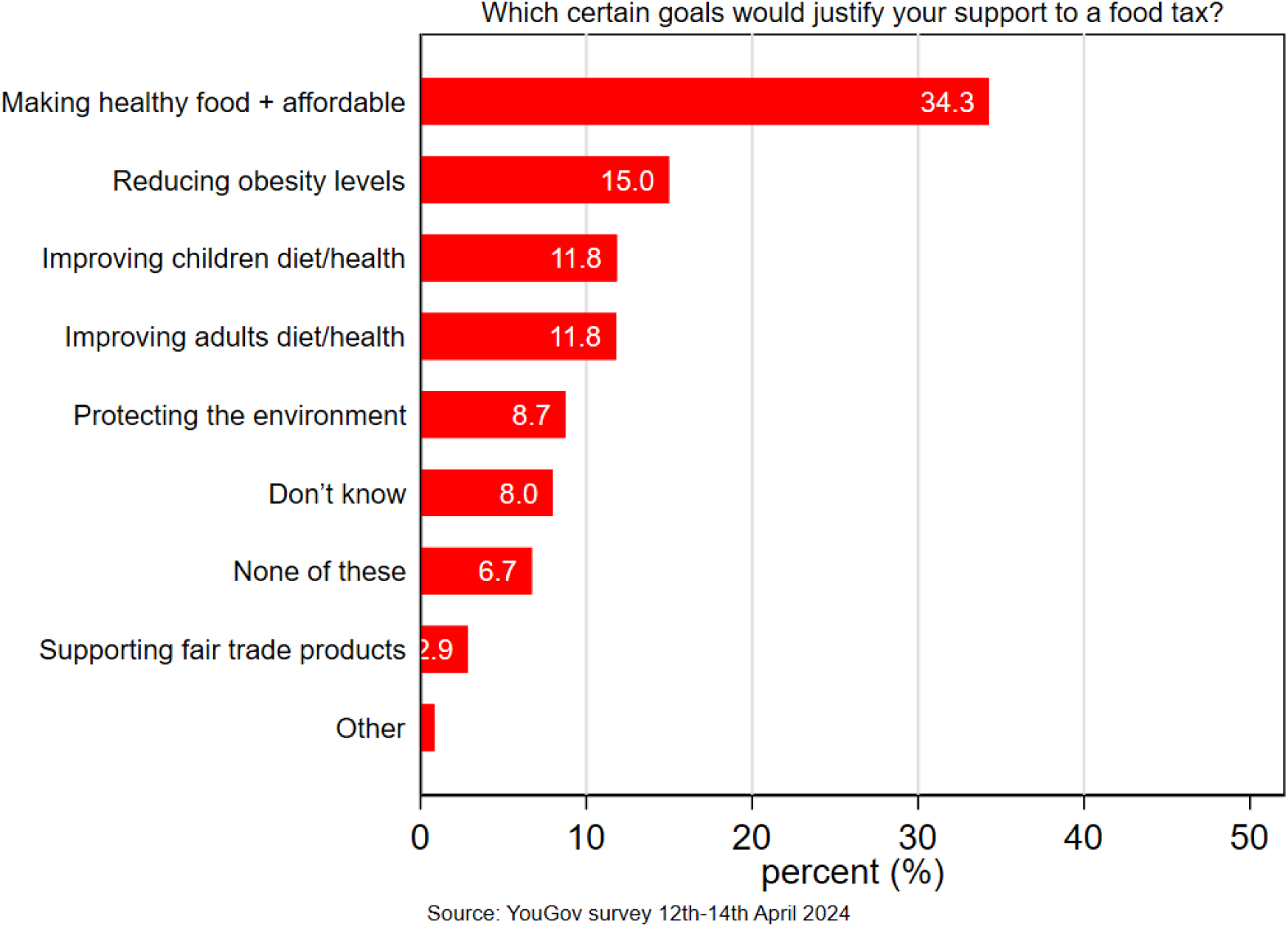
Share of respondents on each of the goals that could be tackled by taxing food and drink products *Notes:* Total sample size are 2,125 adults. Fieldwork was undertaken between 12th - 14th April 2024. Sample weights are used. Question 8: “Some taxes are applied to products to specifically achieve certain goals, either through the revenue raised or by discouraging people from purchasing those products and encouraging them to purchase others instead. Thinking about different goals that could be tackled by taxing food and drink products, which of the following do you think is most important?”.

### 3.3. Insights on preferences for policy design and purpose

Table 2 shows levels of support for a possible tax on unhealthy foods for the full sample and specifically for those who had reported supporting or opposing taxes on unhealthy foods. Overall, we can see a similar pattern of support when comparing the full sample to just those who supported a tax, with higher support and lower opposition seen for specific categories amongst those who supported a tax. A much bigger difference between support and opposition was also seen within the smaller sample of supporters.

**Table 2.** Support for food taxes by food categories and level of support for a tax on unhealthy foods.

| Food category | Full sample<br>(N= 2,125) |  |  | Support a tax<br>(N= 1,047) |  |  | Oppose a tax<br>(N= 924) |  |  | Don't know<sup>±</sup><br>(N=154) |  |  |
| --- | --- | --- | --- | --- | --- | --- | --- | --- | --- | --- | --- | --- |
|  | Support | Oppose | Do not know | Support | Oppose | Do not know | Support | Oppose | Do not know | Support | Oppose | Do not know |
| *Milkshakes* | 32.66 | 52.01 | 15.32 | 51.73 | 35.78 | 12.50 | 15.04 | 73.32 | 11.63 | 15.01 | 32.25 | 52.72 |
| *Fruit juice* | 11.56 | 75.60 | 12.84 | 18.95 | 69.67 | 11.39 | 4.64 | 86.90 | 8.46 | 5.23 | 49.03 | 45.74 |
| *Fresh fruit and vegetables* | 1.37 | 91.41 | 7.22 | 1.72 | 93.98 | 4.51 | 0.86 | 94.37 | 4.77 | 1.06 | 61.84 | 37.10 |
| *Potato crisps* | 40.87 | 45.99 | 13.14 | 65.13 | 24.05 | 9.82 | 18.50 | 71.67 | 9.83 | 18.07 | 30.47 | 51.46 |
| *Red and processed meat* | 22.92 | 64.62 | 12.46 | 37.78 | 51.42 | 10.79 | 8.15 | 83.95 | 7.90 | 14.95 | 37.41 | 47.62 |
| *Cakes* | 44.68 | 41.96 | 13.36 | 68.73 | 21.14 | 10.13 | 22.22 | 67.29 | 10.49 | 23.74 | 27.60 | 48.66 |
| *Ready meals* | 36.20 | 49.96 | 13.84 | 56.55 | 32.31 | 11.14 | 17.38 | 72.64 | 9.98 | 17.39 | 31.18 | 51.43 |
| *Hot takeaways and deliveries* | 39.51 | 47.09 | 13.40 | 60.22 | 29.20 | 10.58 | 20.18 | 69.67 | 10.15 | 21.39 | 30.27 | 48.33 |

Notes: The total sample size is 2,125 adults. Fieldwork was undertaken between 12th - 14th April 2024. These results come from the following question: "Do you think the following foods should or should not have a higher tax applied to them?" and include average sample weights. N= number of observations. <sup>±</sup> 'Don't know' could mean the person does not know/have a view, does not understand the question or does not want to answer the question.

The only category which had more support than opposition amongst the full sample was cake, with 44.68% reporting to support a potential tax on this category (versus 41.96% who opposed it). This figure increased to 68.73% support (and 21.14% opposed) when only looking at those who were generally supportive of a tax. The second most supported category was potato crisps (40.87% support vs 45.99% opposed); however, again, support increased to 65.13% (vs 24.04%) when looking only at those who supported a tax. Other categories with support include hot takeaways (39.51%) and ready meals (36.20%). Support was low for a tax on red and processed meat (22.92%) and, unsurprisingly, fruit and vegetables (1.37%). Overall, we can see more support for a tax on discretionary products which fall outside of the Government recommended Eatwell Plate (e.g. cakes and biscuits) rather than meals (ready meals and takeaways) and less processed foods (e.g. fruits, vegetables and meat).

To better understand what criteria/outcomes respondents most value to justify a tax on foods, respondents were asked about their priority goal that a tax should address. The highest support of the options provided was for making healthier food more affordable (34.3%), followed by obesity (15%) and improving children’s and adults’ diet and health (both at 11.8%). Fewer people put fair trade and environmental outcomes as their priority. The high support for a tax which can make healthier food more affordable is consistent with the findings that there is greater support for a tax if it is also used to make healthier food more affordable. It is also reflective of the current cost-of-living crisis, which has led to 1 in 5 households reporting to be affected by food insecurity in January 2024 (Food Foundation, 2024) and food prices increasing by roughly 25% (Food Foundation, 2024).

## 4. DISCUSSION

Our representative national survey of 2,125 people in the United Kingdom provides valuable insights into public attitudes towards food taxes aimed at promoting healthier eating. The findings highlight several key aspects that policymakers need to consider when designing and implementing such taxes.

While the survey shows a considerable degree of support for the idea of taxing unhealthy foods, the extent of this support varies across different groups of respondents. Support for the idea of a tax is also significantly higher when it is coupled with efforts to make healthier food more affordable or cheaper. The survey also reveals that a substantial share of respondents had poor knowledge about taxes currently applied on food and non-alcoholic beverages in the UK. This lack of awareness and knowledge about the health impacts of unhealthy foods and the potential benefits of food taxes may play an important role in shaping public attitudes towards taxes. Respondents expressed varying preferences regarding the specific foods to be taxed, goals and the allocation of tax revenues. Importantly, there is a preference for taxing discretionary items such as cakes and crisps rather than meals and for revenues to help make healthier food more affordable. In addition, the most supported goal of a food tax is to make healthier foods more affordable. Understanding these preferences can help policymakers design food taxes that are more likely to gain public support and achieve desired health outcomes.

In conclusion, the survey provides evidence that carefully designed food taxes aimed at incentivising healthy eating and curbing diet-related diseases can find support from a sizable majority of the UK population, provided that attention is paid to the affordability of healthy foods in designing the tax incentives.

## Data Availability

All data produced in the present study are available upon reasonable request to the authors. Data may be obtained from a third party (YouGov Plc.) and are not publicly available.

## Acknowledgements

This study forms part of the Fiscal INCentives for Health improvement: repurposing consumption taxes on food (FINCH) project, funded through the NIHR Public Health Research Programme (NIHR133974). The views expressed are those of the author(s) and not necessarily those of the NIHR or the Department of Health and Social Care.

## Data sharing statement

Data may be obtained from a third party (YouGov Plc.) and are not publicly available.

## Ethics Approval Statement

Name of Committee: Imperial College London Research Governance and Integrity Team Reference no: 22IC7545

## SUPPLEMENTARY APPENDIX

### APPENDIX A. Further Tables and Figures

**Table A1.** Descriptive statistics for sample party policy preferences.

|  |  | Weighted Sample |  | Unweighted Sample |  | Population |
| --- | --- | --- | --- | --- | --- | --- |
|  |  | N | % | N | % | % |
| Vote in 2019<br>General Elections | Con | 678 | 31.9 | 690 | 32.5 | 43.6 |
|  | Lab | 497 | 23.4 | 494 | 23.2 | 32.1 |
|  | Lib Dem | 178 | 8.4 | 179 | 8.4 | 11.5 |
|  | Do not know/skipped |  |  | 553 | 26 |  |
| EU Referendum<br>2016 | Remain | 746 | 35.1 | 861 | 40.5 | 48.11 |
|  | Leave | 788 | 37.1 | 775 | 36.5 | 51.89 |
|  | Did not vote/can't remember |  |  | 489 | 23 |  |

*Notes:* The total sample size is 2,125 adults. Fieldwork was undertaken between 12th - 14th April 2024. The survey was carried out online. The figures have been weighted and are representative of all UK adults (aged 18+). *Source:* YouGov and Office National Statistics (ONS).

**Figure A1.**
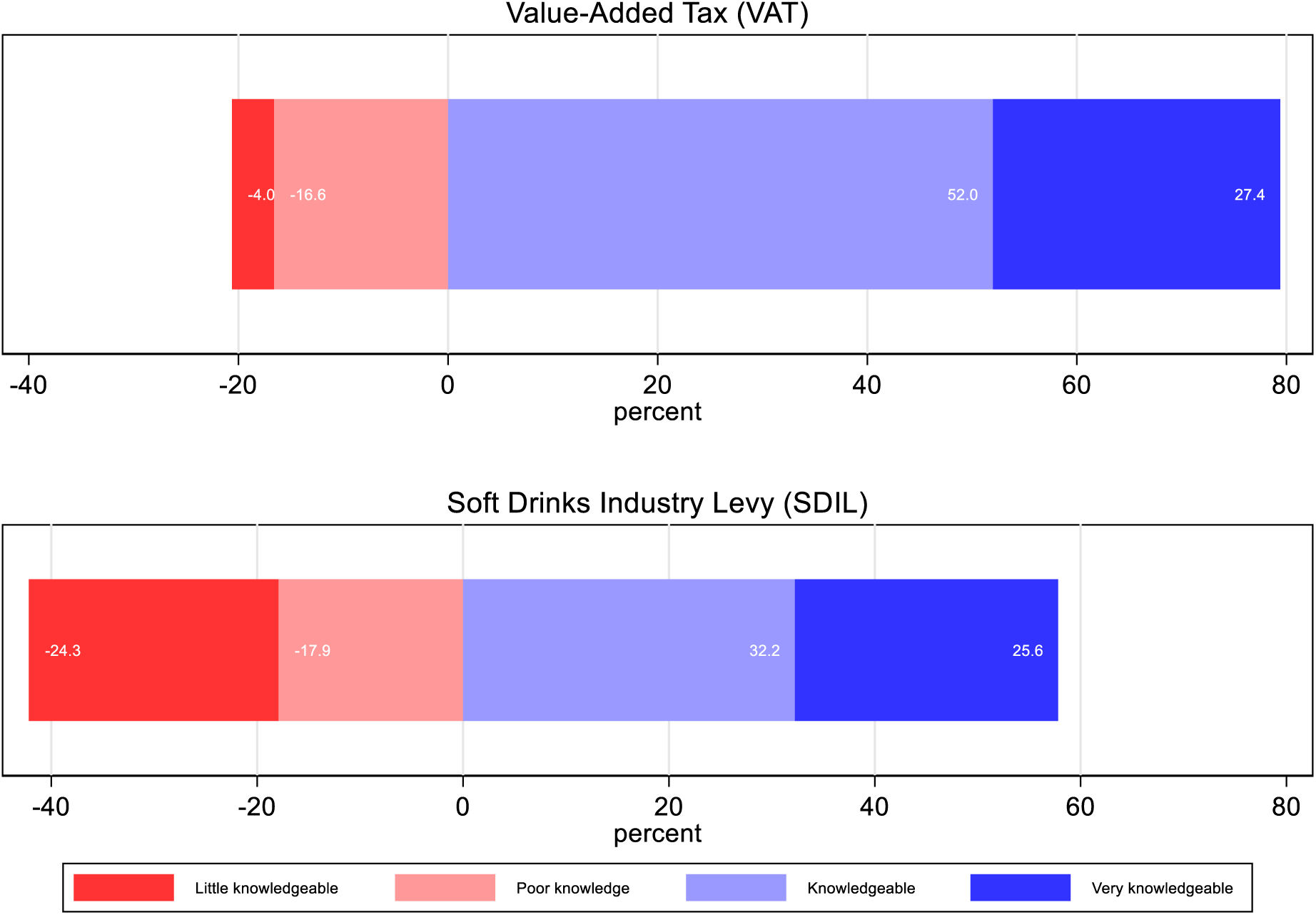
Self-reported knowledge of taxes on food and beverages in the United Kingdom: share of respondents. *Notes*: Notes: Total sample size are 2,125 adults. Fieldwork was undertaken between 12th - 14th April 2024. Self-reported knowledge of taxes on food and beverages are defined as follows: poor knowledge: “I have never heard of it and don’t know anything about what foods it is added to”; Little knowledgeable: “I have heard of it, but don’t know anything about what foods it is added to”; Knowledgeable: “I know what it is, but know little about what foods it is added to”; and Very knowledgeable: “I know what it is, and the foods it is added to”. We are using average weights. Sample weights are used.

**Figure A2.**
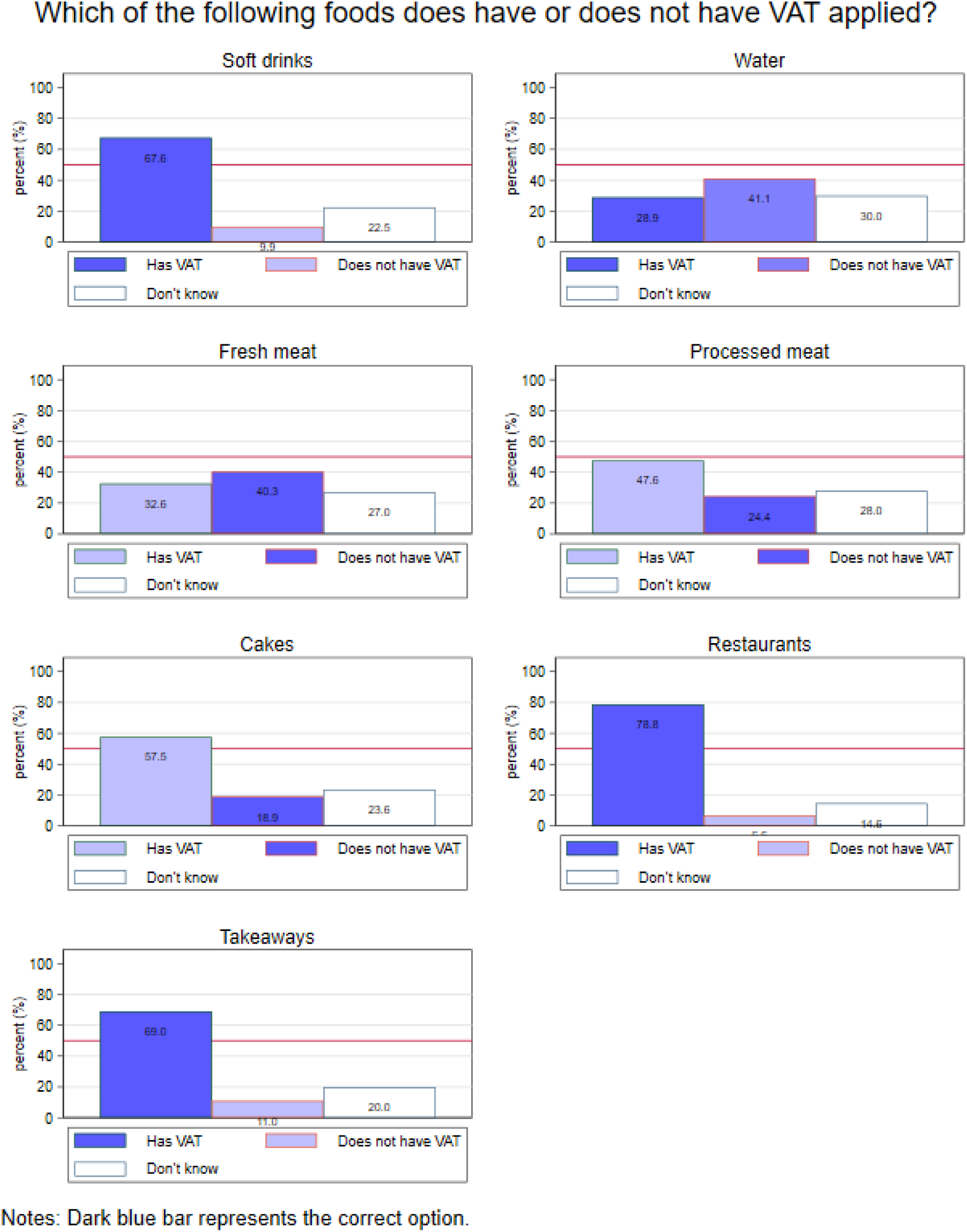
Knowledge about food groups which have VAT applied: Share of correct answers in dark blue. *Notes:* The total sample size is 2,125 adults. Fieldwork was undertaken between 12th - 14th April 2024. Sample weights are used.

**Figure A3.**
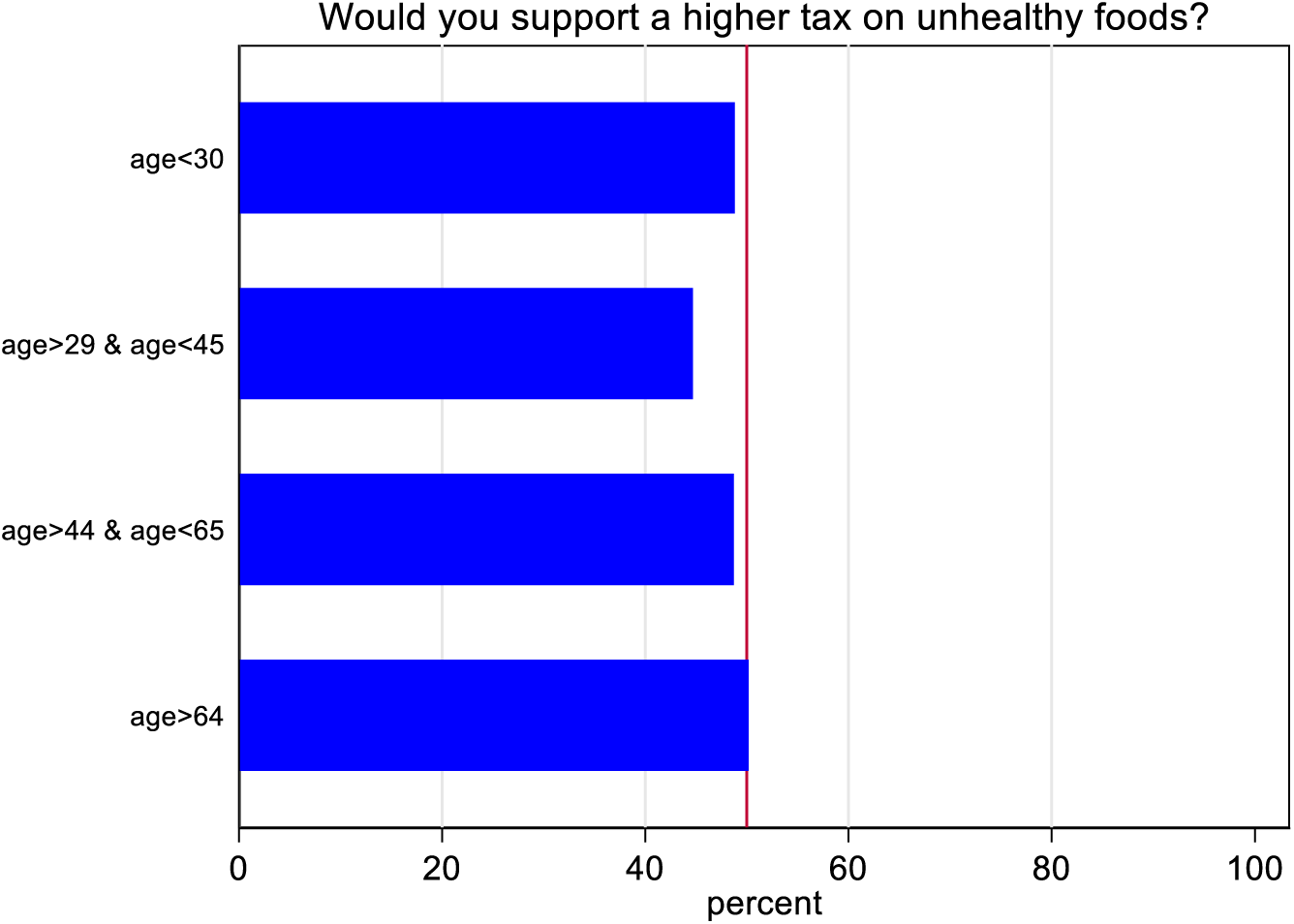
Share of respondents support a higher tax on unhealthy foods by age groups. *Notes:* The total sample size is 2,125 adults. Fieldwork was undertaken between 12th - 14th April 2024. The base category includes the following responses: Strongly/somewhat opposed/Don’t know. The support category includes responses that Strongly/somewhat support. Sample weights are used.

**Figure A4.**
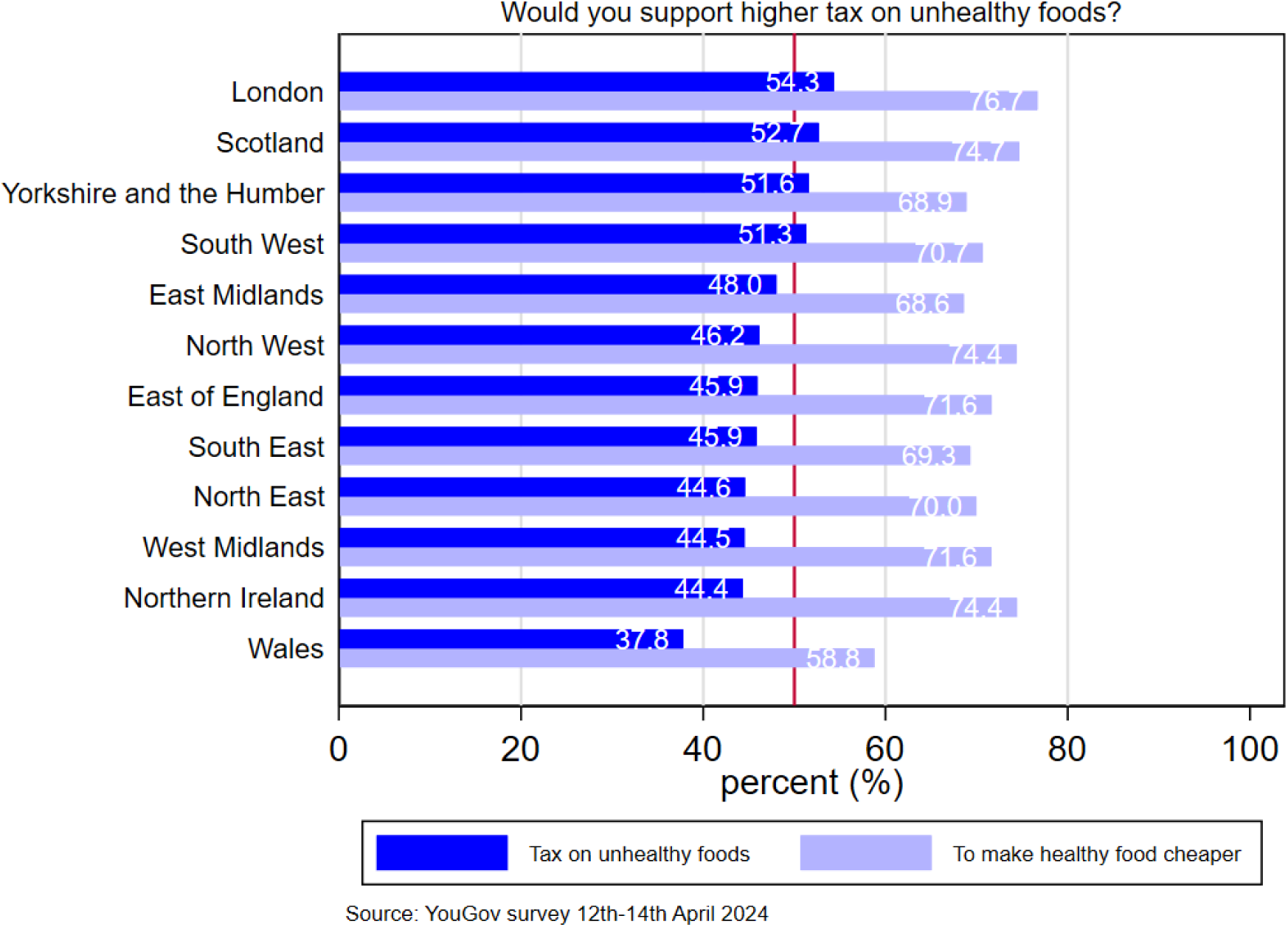
Share of respondents who agree (somewhat to strongly) that “would you support a higher tax on unhealthy foods?” or “Would you support a higher tax on unhealthy foods to make healthy food cheaper” by regions. Notes: The total sample size is 2,125 adults; the base category includes the following responses: Strongly/somewhat opposed/Don’t know. The support category includes responses that Strongly/somewhat support. Sample weights are used.

**Figure A5.**
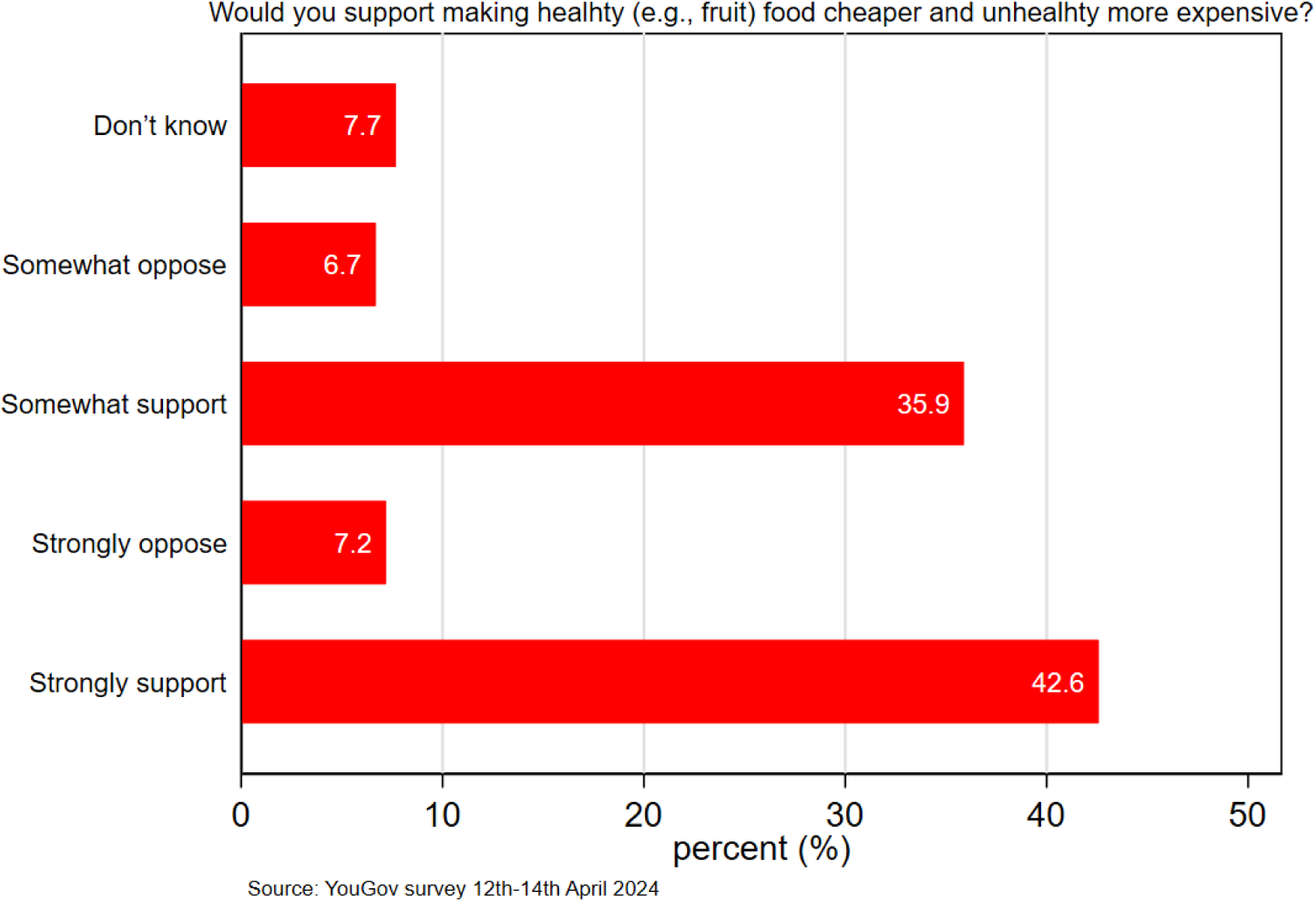
Support for tax on unhealthy foods between different food categories. Notes: The total sample size is 2,125 adults. Sample weights are used.

**Figure A6.**
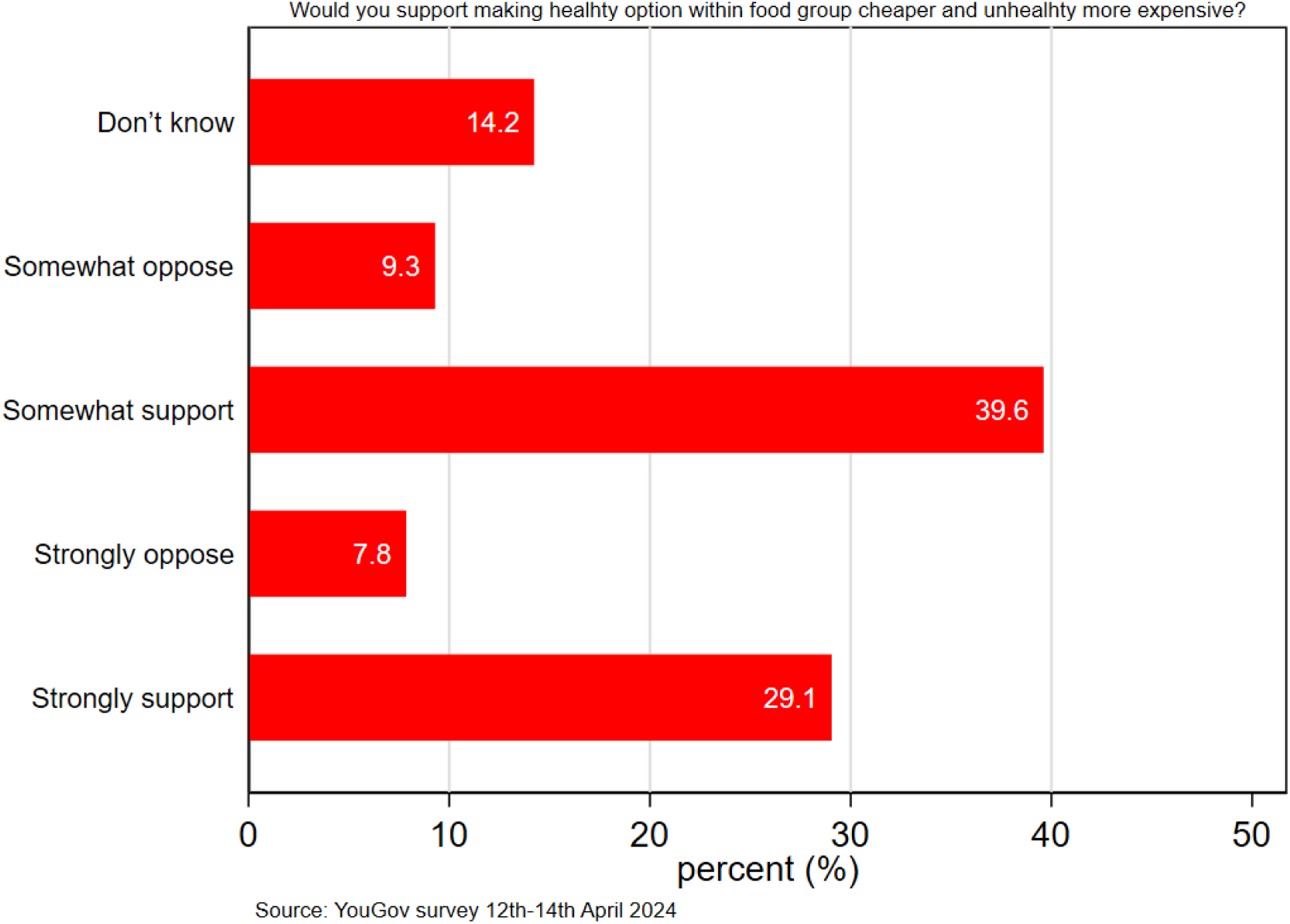
Support for tax on unhealthy foods within foods in the same category. Notes: The total sample size is 2,125 adults. Sample weights are used.

### APPENDIX B. Empirical Methodology and Robustness Checks

To investigate the relationship between individuals’ sociodemographic characteristics and support for unhealthy food taxes, we estimate the following regression:

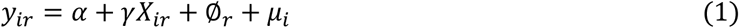

Where *y*_*i*_ is a set of outcome variables capturing to what extent support a tax on unhealthy foods by person *i*. These variables are binary regarding the extent of support for taxes on unhealthy food, taking values one if the respondent *”strongly/somewhat supports”* and zero if the respondent *”strongly/somewhat opposes.”* Sociodemographic characteristics included in the analysis are age groups, a binary variable capturing high social class (takes one if the respondent’s social class is *ABC1*, and zero if *C2DE.* We also include political options in the last 2019 election and region/country fixed effects. We estimate both a linear probability model (LPM) and a logit fixed-effect model (Logit-FE). We provide estimates with robust standard errors and survey weights are used in all the regressions.

The following robustness checks have been performed, including (i) a hypothesis test on the statistical differences among the control variables for our two main outcomes, (ii) a multicollinearity test, Variance inflation factor (VIF), and correlation plots, and (iii) logistic regressions as a main econometric specification.

**1. Test for supporting tax on subsidy:**

~~~
. test age_cat_2 age_cat_3 age_cat_4 gender highsocial_class London England
(1) age_cat_2 = 0
(2) age_cat_3 = 0
(3) age_cat_4 = 0
(4) gender = 0
(5) highsocial_class = 0
(6) London = 0
(7) England = 0
     F(7, 1987) = 7.64
        Prob > F = 0.0000
~~~

**2. Test for supporting taxon unhealthy foods**

~~~
. test age_cat_2 age_cat_3 age_cat_4 gender highsocial_class London England
(1) age_cat_2 = 0
(2) age_cat_3 = 0
(3) age_cat_4 = 0
(4) gender = 0
(5) highsocial_class = 0
(6) London = 0
(7) England = 0
     F(7, 1963) = 4.37
        Prob > F = 0.0001
~~~

**Table B1.**
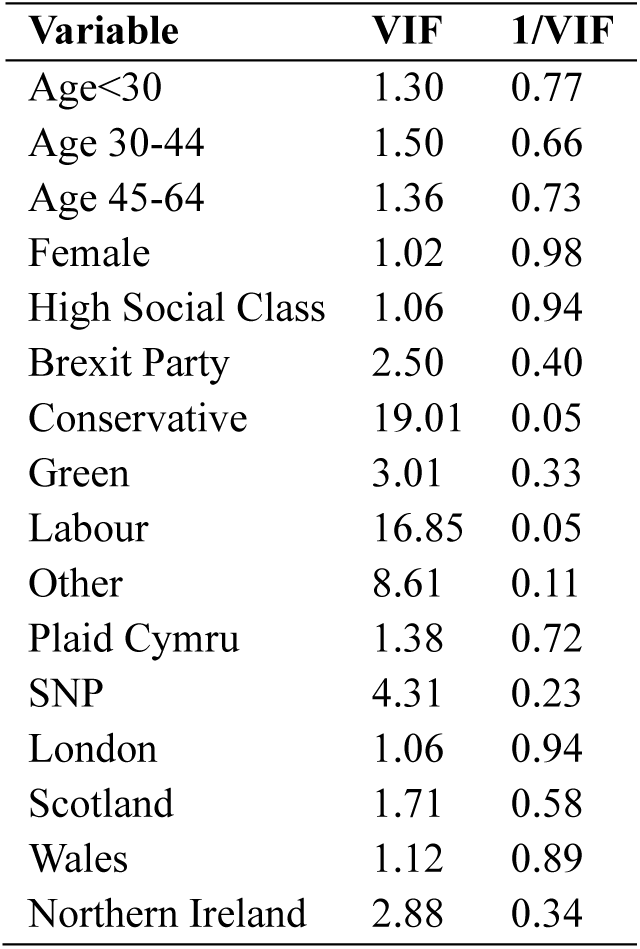
Multicollinearity test (VIF) among socioeconomic variables.

To interpret the results:

- A value of 1 indicates there is no correlation between a given explanatory variable and any other explanatory variables in the model.
- A value between 1 and 5 indicates moderate correlation between a given explanatory variable and other explanatory variables in the model, but this is often not severe enough to require attention.
- A value greater than 5 indicates potentially severe correlation between a given explanatory variable and other explanatory variables in the model. In this case, the coefficient estimates and p-values in the regression output are likely unreliable.

**Figure B1.**
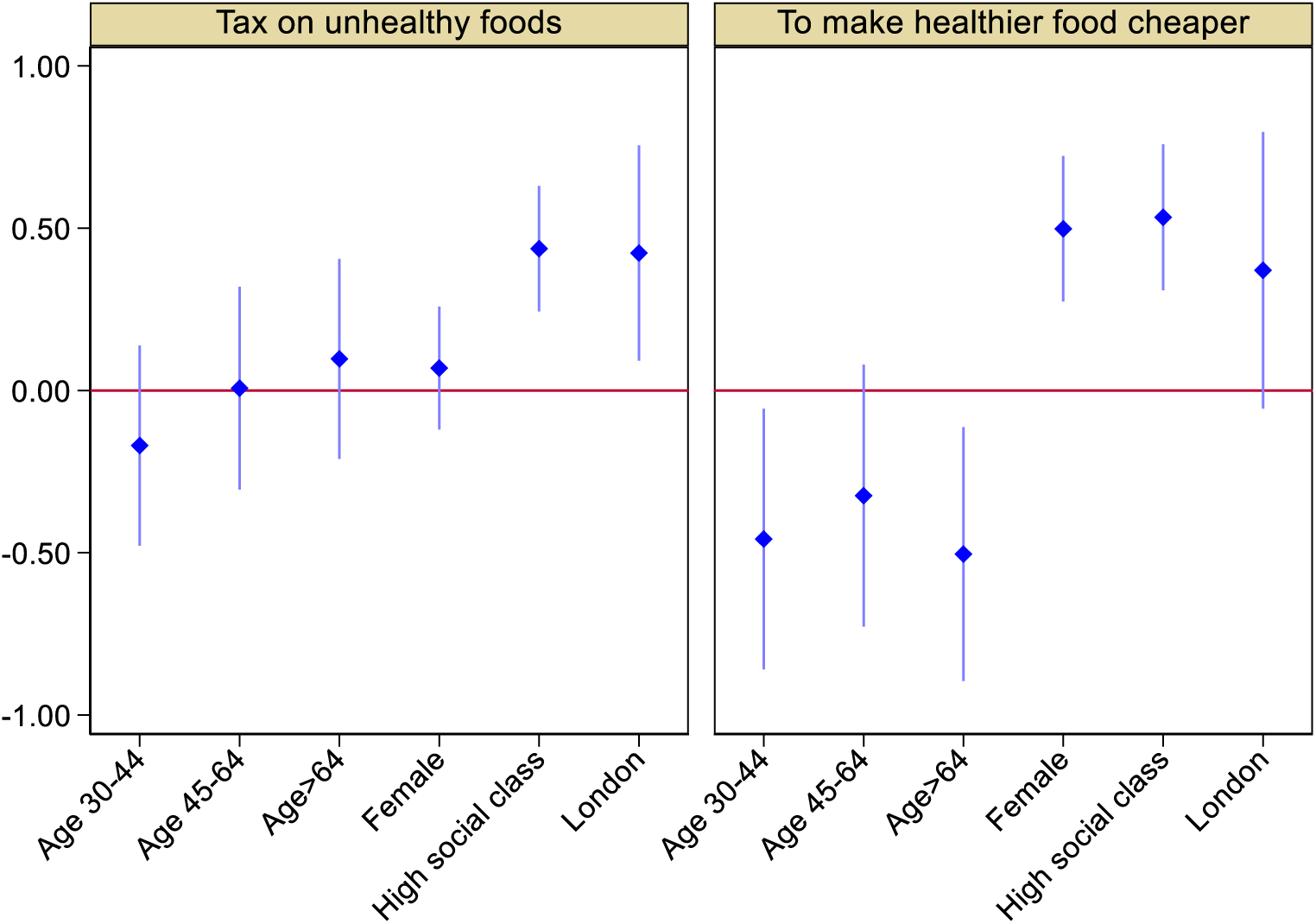
Logistic regression results for our main two outcomes on tax support on unhealthy foods. Notes: The total sample size is 1,971. adults. ‘Do not know’ observations are not included in the analysis. Sample weights are used. The age case category is age>30.

### Appendix C. The Questionnaire and variable definition

{single}

**Q1) Generally speaking, would you support or oppose a higher tax on unhealthy foods?**

<1> Strongly support
<2> Somewhat support
<3> Somewhat oppose
<4> Strongly oppose
<5> Don’t know

{single}

**Q2) And would you support or oppose a higher tax on unhealthy foods if the money raised was used directly to help make healthier food cheaper?**

<1> Strongly support
<2> Somewhat support
<3> Somewhat oppose
<4> Strongly oppose
<5> Don’t know

[grid]

**Q3) How much, if at all, do you know about the following taxes and which foods they are added to?**

- **Value Added Tax (VAT)**
- **Soft Drink Industry Levy (SDIL)**
<1> I know what it is, and the foods it is added to
<2> I know what it is, but know little about what foods it is added to
<3> I have heard of it, but don’t know anything about what foods it is added to
<4> I have never heard of it and don’t know anything about what foods it is added to

[grid]

**Q4) Which of the following foods do you think have and don’t have VAT added to them?**

A. Soft drinks
B. Bottled water
C. Fresh meat
D. Processed meat (e.g. ham)
E. Cakes
F. Chocolate and confectionery
G. Food eaten in restaurants and cafes
H. Hot takeaways and deliveries
<1> Does have VAT
<2> Does not have VAT
<3> Don’t know
**Q4) Which of the following drinks do you think have and don’t have Soft Drinks Industry Levy (SDIL) added to them?**

A. Soft drinks
B. Bottled water
C. Fruit juice
D. Milkshakes
<1> Does have SDIL
<2> Does not have SDIL
<3> Don’t know

[grid]

**5) Do you think the following foods should or should not have a higher tax applied to them?**

A. Milkshakes
B. Fruit juice
C. Fresh fruit and vegetables
D. Potato crisps
E. Red and processed meat
F. Cakes
G. Ready meals
H. Hot takeaways and deliveries
<1> Should have a higher tax
<2> Should not have a higher tax
<3> Don’t know

{Single}

**Q6) Some taxes are applied to products to specifically achieve certain goals either through the revenue raised or by discouraging people to purchase those products and encouraging them to purchase others instead.**

**Thinking about different goals that could be tackled by taxing food and drink products, which of the following do you think are most important?**

<1> Protecting the environment

<2> Improving adults’ diet and health

<3> Supporting fair trade products

<4> Making healthy food more affordable

<6> Improving children’s diet and health

<7> Reducing obesity levels

<8> Other

<9> None of these

<10> Don’t know

{grid}

**Q7) Would you support or oppose the following…**

- Making it less expensive for people to choose healthy foods (e.g. fruit and vegetables), and more expensive to choose less healthy foods (e.g. deserts, prepared meals)
- Making it less expensive for people to choose healthier options within food groups (e.g. ready meals containing less fat, salt and sugar) and more expensive to choose less healthy options within the same food group
<1> Strongly support
<2> Somewhat support
<3> Strongly oppose
<4> Somewhat oppose
<5> Don’t know

**Table C1.**
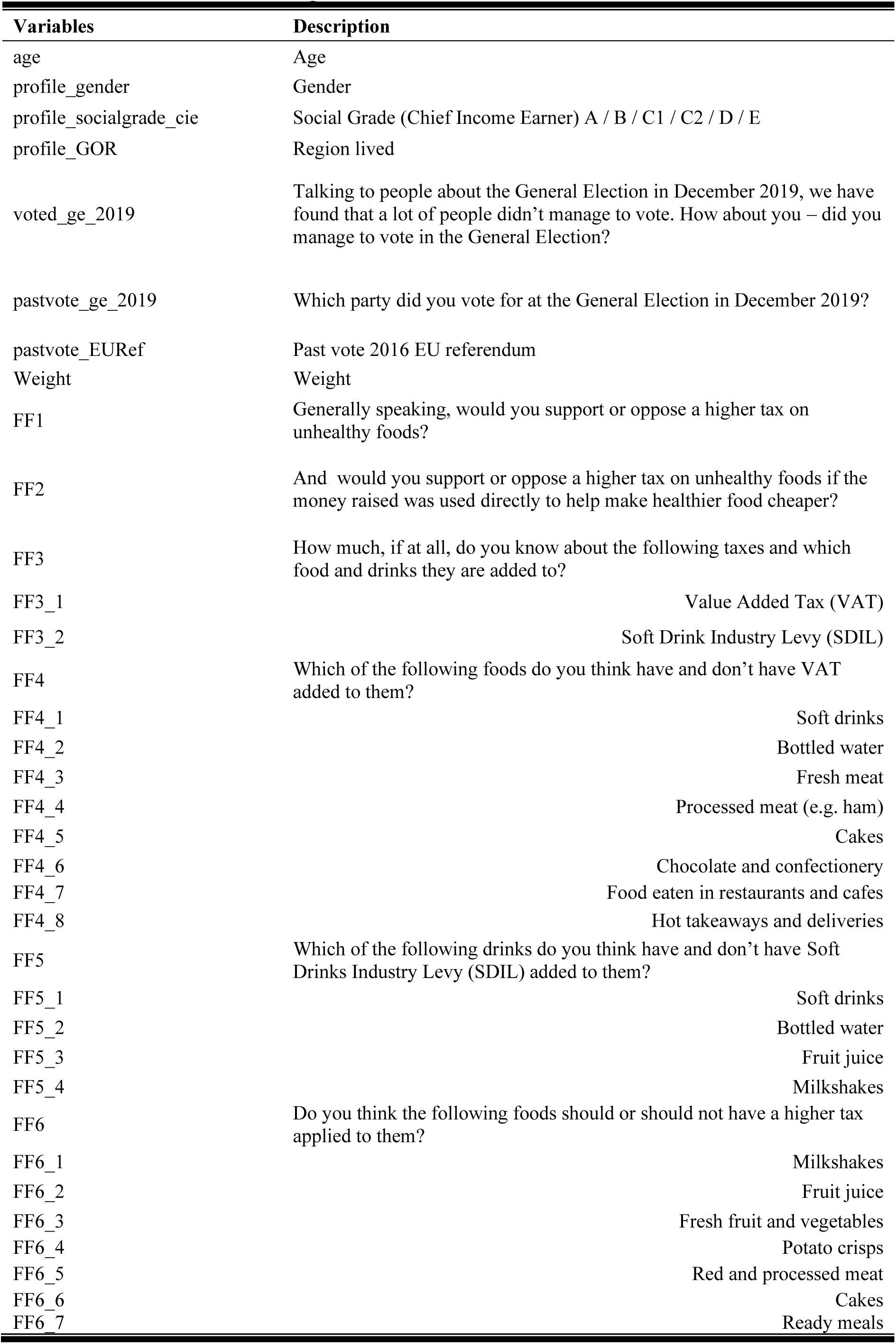
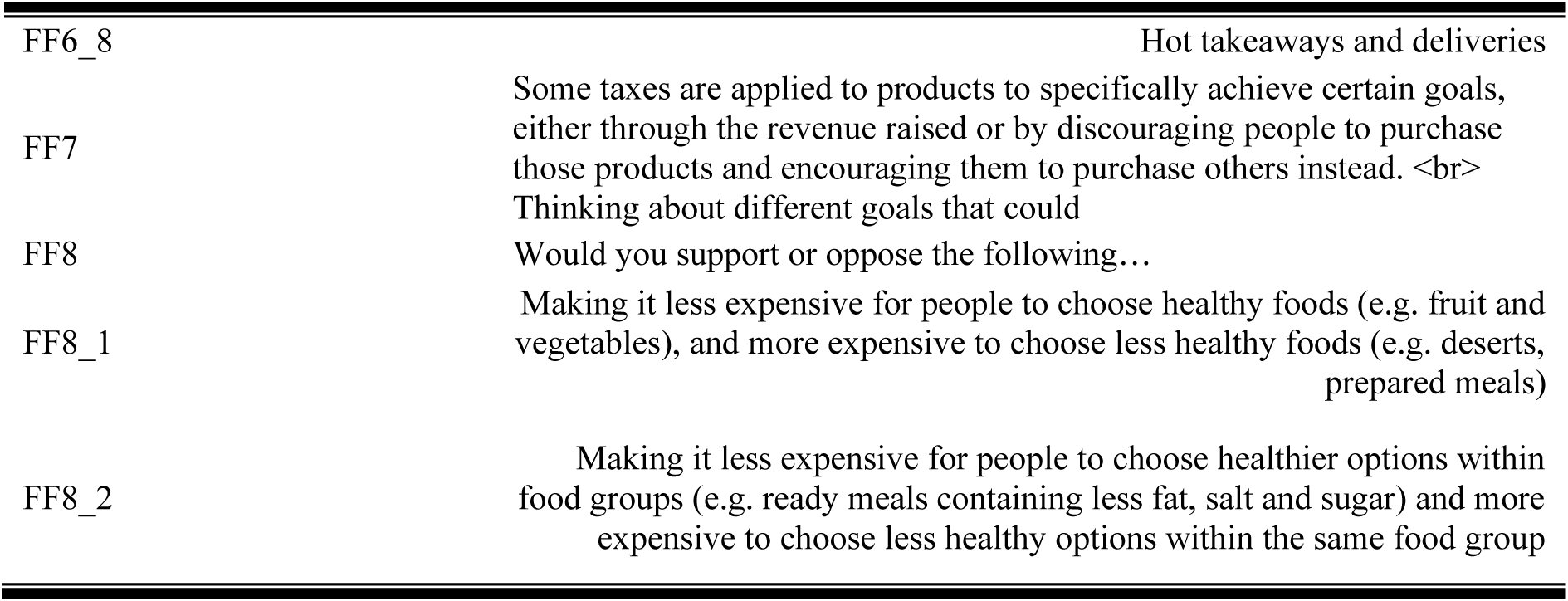
YouGov variables description.

| Variables | Description |
| --- | --- |
| age | Age |
| profile_gender | Gender |
| profile_socialgrade_cie | Social Grade (Chief Income Earner) A / B / C1 / C2 / D / E |
| profile_GOR | Region lived |
| voted_ge_2019 | Talking to people about the General Election in December 2019, we have found that a lot of people didn't manage to vote. How about you – did you manage to vote in the General Election? |
| pastvote_ge_2019 | Which party did you vote for at the General Election in December 2019? |
| pastvote_EURef | Past vote 2016 EU referendum |
| Weight | Weight |
| FF1 | Generally speaking, would you support or oppose a higher tax on unhealthy foods? |
| FF2 | And would you support or oppose a higher tax on unhealthy foods if the money raised was used directly to help make healthier food cheaper? |
| FF3 | How much, if at all, do you know about the following taxes and which food and drinks they are added to? |
| FF3_1 | Value Added Tax (VAT) |
| FF3_2 | Soft Drink Industry Levy (SDIL) |
| FF4 | Which of the following foods do you think have and don't have VAT added to them? |
| FF4_1 | Soft drinks |
| FF4_2 | Bottled water |
| FF4_3 | Fresh meat |
| FF4_4 | Processed meat (e.g. ham) |
| FF4_5 | Cakes |
| FF4_6 | Chocolate and confectionery |
| FF4_7 | Food eaten in restaurants and cafes |
| FF4_8 | Hot takeaways and deliveries |
| FF5 | Which of the following drinks do you think have and don't have Soft Drinks Industry Levy (SDIL) added to them? |
| FF5_1 | Soft drinks |
| FF5_2 | Bottled water |
| FF5_3 | Fruit juice |
| FF5_4 | Milkshakes |
| FF6 | Do you think the following foods should or should not have a higher tax applied to them? |
| FF6_1 | Milkshakes |
| FF6_2 | Fruit juice |
| FF6_3 | Fresh fruit and vegetables |
| FF6_4 | Potato crisps |
| FF6_5 | Red and processed meat |
| FF6_6 | Cakes |
| FF6_7 | Ready meals |
| FF6_8 | Hot takeaways and deliveries |
| FF7 | Some taxes are applied to products to specifically achieve certain goals, either through the revenue raised or by discouraging people to purchase those products and encouraging them to purchase others instead. <br>Thinking about different goals that could |
| FF8 | Would you support or oppose the following... |
| FF8_1 | Making it less expensive for people to choose healthy foods (e.g. fruit and vegetables), and more expensive to choose less healthy foods (e.g. deserts, prepared meals) |
| FF8_2 | Making it less expensive for people to choose healthier options within food groups (e.g. ready meals containing less fat, salt and sugar) and more expensive to choose less healthy options within the same food group |

